# Evaluation of ChatGPT’s Pathology Knowledge using Board-Style Questions

**DOI:** 10.1101/2023.10.01.23296400

**Authors:** Saroja D Geetha, Anam Khan, Atif Khan, Bijun S Kannadath, Taisia Vitkovski

## Abstract

**Objectives:** ChatGPT is an artificial intelligence (AI) chatbot developed by OpenAI. Its extensive knowledge and unique interactive capabilities enable it to be utilized in various innovative ways in the medical field such as writing clinical notes, simplifying radiology reports. Through this study we aim to analyze its pathology knowledge to advocate its role in transforming pathology education.

**Methods:** American Society for Clinical Pathology (ASCP) Resident Question bank (RQB) 2022 was used to test ChatGPT v4. Practice tests were created in each sub-category and were answered based on the input provided by ChatGPT. Questions that required interpretation of images were excluded. ChatGPT’s performance was analyzed and compared with the average peer performance.

**Results:** The overall performance of ChatGPT was 56.98%, lower than that of the average peer performance of 62.81%. ChatGPT performed better on clinical pathology (60.42%) than anatomic pathology (54.94%). Furthermore, its performance was better on easy questions (68.47%) compared to intermediate (52.88%) and difficult questions (37.21%).

**Conclusions:** ChatGPT has the potential to be a valuable resource in pathology education if trained on a larger, specialized medical dataset. Relying on it solely for the purpose of pathology training should be with caution, in its current form.

**Key points:**

- ChatGPT is an AI chatbot, that has gained tremendous popularity in multiple industries, including healthcare. We aim to understand its role in revolutionizing pathology education.
- We found that ChatGPT’s overall performance in Pathology Practice Tests were lower than that expected from an AI tool, furthermore its performance was subpar compared to pathology residents in training.
- In its current form ChatGPT is not a reliable tool for pathology education, but with further refinement and training it has the potential of being a learning asset.

## Introduction

ChatGPT (Generative Pretrained Transformer) is an Artificial intelligence (AI) chatbot developed by OpenAI. It is a large language model (LLM) and as the name suggests it is highly pretrained on large data sets to generate textual replies and solutions when users converse with it based on a neural network that works on the transformer architecture.^1^ First introduced in 2018 and trained on a dataset of 40GB, it has undergone multiple revisions and updates. In 2020, GPT-3 that was trained on a much larger dataset of 570GB, with improved capabilities in generating human-like responses for interactive conversations was released.^2^ ChatGPT-4, the latest version, further builds upon GPT-3 to provide more natural and engaging dialogue experiences with users.

ChatGPT demonstrates a high level of user-friendliness and is accessible to users without any cost and offers prompt assistance in a matter of minutes. Its widespread adoption ranges from school students to scientific researchers, vouching for its popularity. Because of its vast vocabulary and capacity to comprehend large knowledge it is capable of writing essays, poems, manuscripts, provide templates for letters, emails and presentation and aid in program coding tasks. ^3^ Additionally, it possesses the ability to adapt and refine written material to align with specific artistic styles. A noteworthy feature of the tool is its capacity to regenerate responses, enabling users to iterate and refine their output if dissatisfied with the initial results.

It proves to be a promising tool that can revolutionize multiple industries, including health care.^4^ It has the potential to make writing clinical notes easier. A recent study also showed that it can effectively simplify and generate radiology reports.^5^ Though it is not trained to be used in medical education, its ability to comprehend large knowledge makes it a possible candidate for enhancing medical education.^6^ Pathology is one of the fore runner fields in medicine to adopt and utilize AI. Through interactive chats ChatGPT has the potential to help medical students, residents, and pathologists to understand complex concepts, such as the pathophysiology of a disease, the histologic features of a specific entity, interpretation of laboratory test results and immunostain panels.^7^ However, the extent to which it can effectively aid in mastering pathology concepts remains an open question. Therefore, we aim to analyze ChatGPT’s pathology knowledge to help advise its role in transforming pathology education.

## Methods

American Society for Clinical Pathology (ASCP) Resident Question bank (RQB) 2022^8^ was used to test ChatGPT. The RQB consists of 6 courses and is subdivided by specialty. The practice tests are customizable and consist of 565 anatomic pathology (AP) and 151 clinical pathology (CP) questions (as of 05/26/2022). The questions in this assessment are of the multiple-choice format. Each question is accompanied by five possible answer choices, with only one of them being correct. Practice tests were created in each sub-category. Questions along with answer stem was put in to ChatGPT v4.^9^ ChatGPT provided us with the answer option (Figure 1). The answers were collected, and the test was answered based on this input. All questions that required interpretation of pictures or images, charts and tables were excluded. Upon completion of each question, ChatGPT’s performance and the percentage of peers who provided the correct answer were recorded. The peer cohort was ascertained by ASCP and encompassed individuals who took the RQB assessment. Since the RQB is designed for pathology board preparation, we believe that the majority of the test takers are residents-in training. The performance of ChatGPT was evaluated both across the entire question bank and within each specific sub-category. Its performance was also assessed in relation to the performance of peers. Based on the peer performance the questions were categorized as easy (>70%), intermediate (40-70%) and difficult (<40%). In addition, to investigate the reproducibility of its responses, a second round was conducted using the same set of questions, and the responses were compared to those obtained in the first test. This is primarily a descriptive study; however statistical analysis was done to compare ChatGPTs performance with that of peer average. Fisher’s exact test was done using Stata Statistical Software: Release 16.1. College Station, TX. This study was exempt from IRB review.

**Figure 1:**
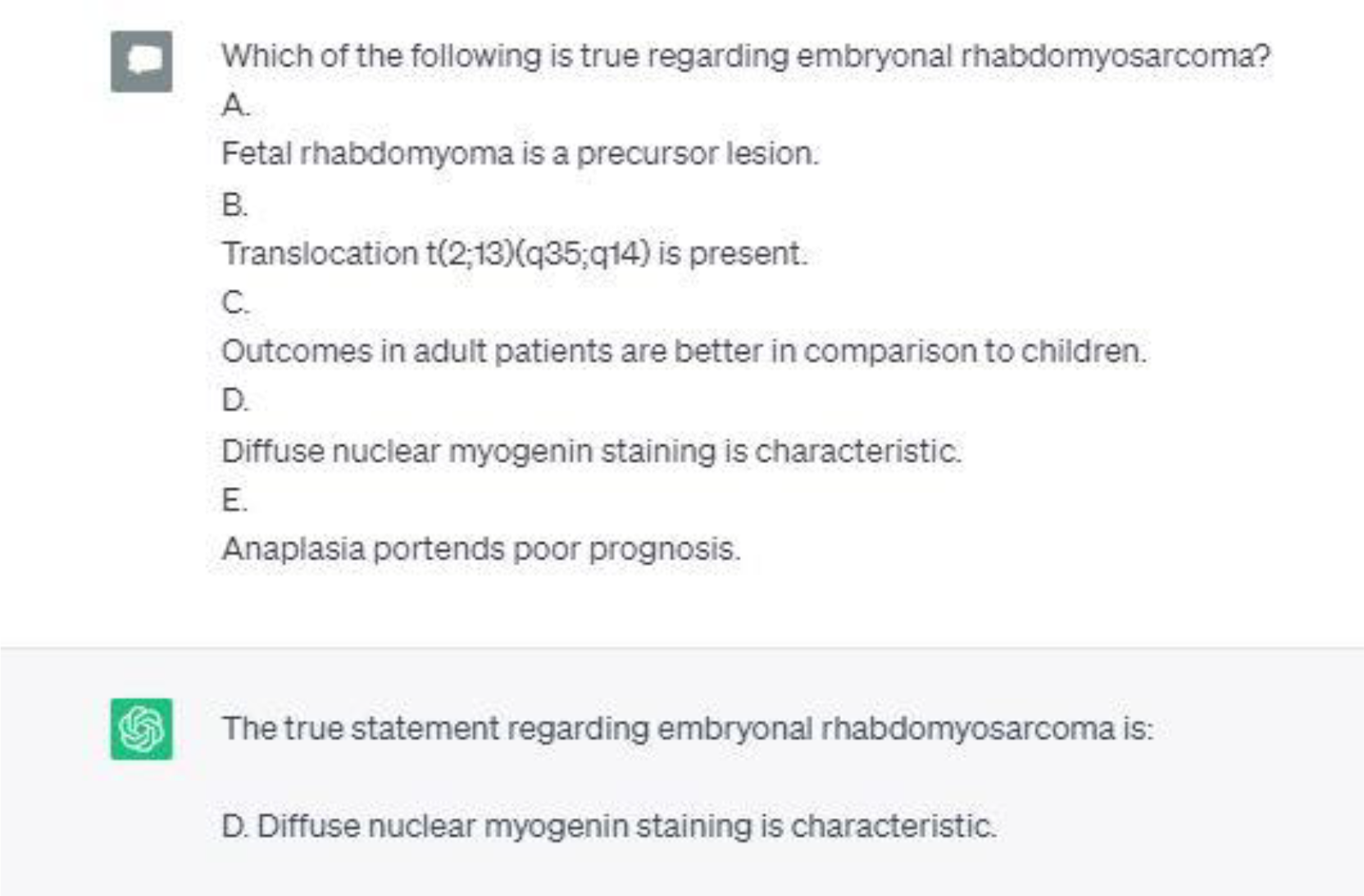
ChatGPT window demonstrating the input (on the upper half), which is the question stem along with the multiple-choice answers fed by our study group, followed by output (on the lower half), which is the answer choice provided by ChatGPT.

## Results

A total of 258 questions that met the inclusion criteria were tested. Of them 162 questions were AP and 96 were CP. Classification based on difficulty level yielded 111 easy, 104 intermediate and 43 difficult questions. The overall performance of ChatGPT was 56.98% (Figure 2) and it was lower than that of the average peer performance (62.81%) (Figure 3). However, this was not statistically significant (Fisher’s exact = 0.209). ChatGPT performed better on the CP section (60.42%) than the AP section (54.94%) (Figure 2). Furthermore, its performance was better on easy questions (68.47%) compared to intermediate (52.88%) and difficult questions (37.21%) (Table 1 and Figure 4).

**Figure 2:**
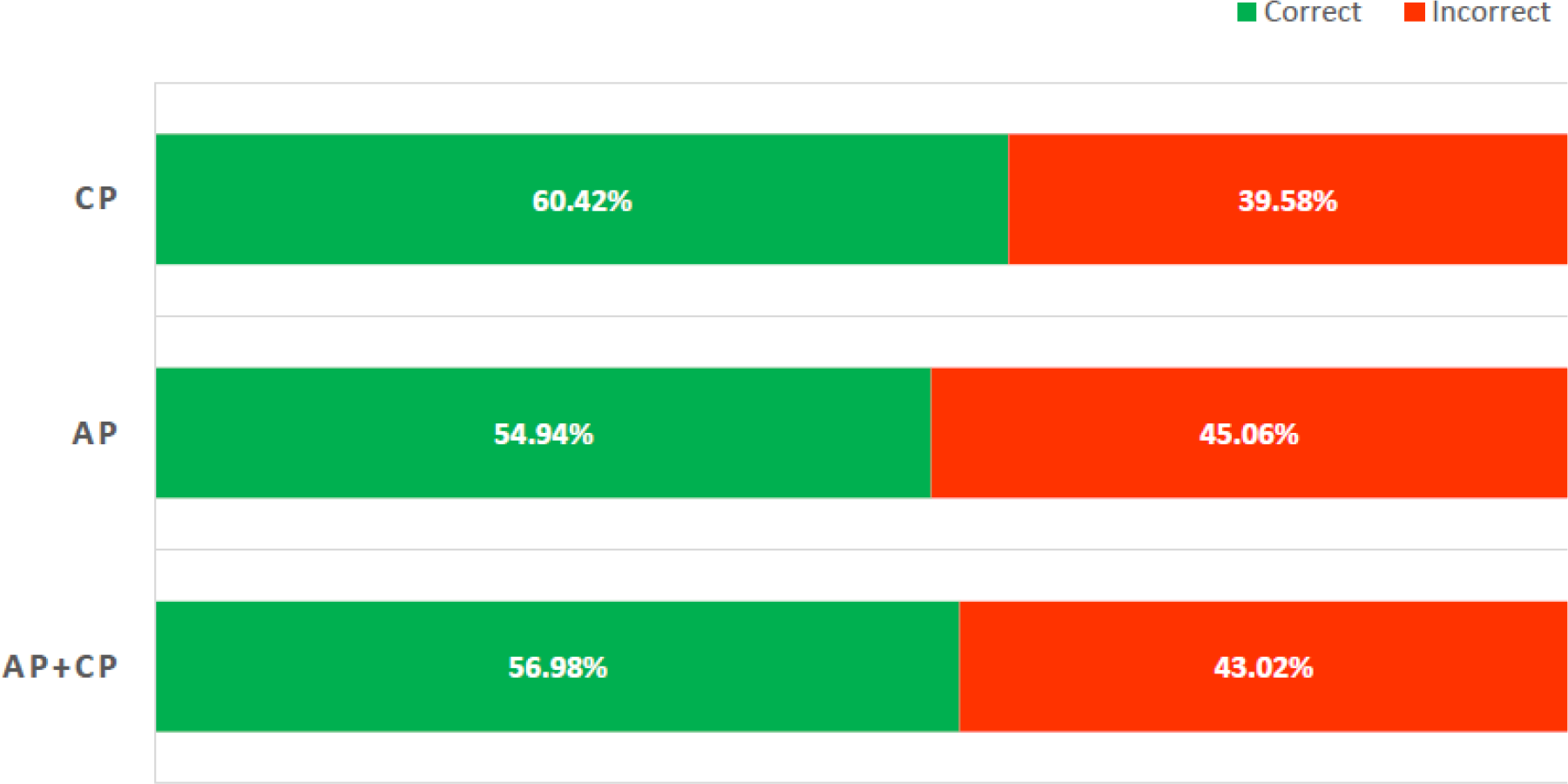
ChatGPTs performance in the ASCP Resident Question Bank 2022. AP: Anatomic pathology; CP: Clinical pathology

**Figure 3:**
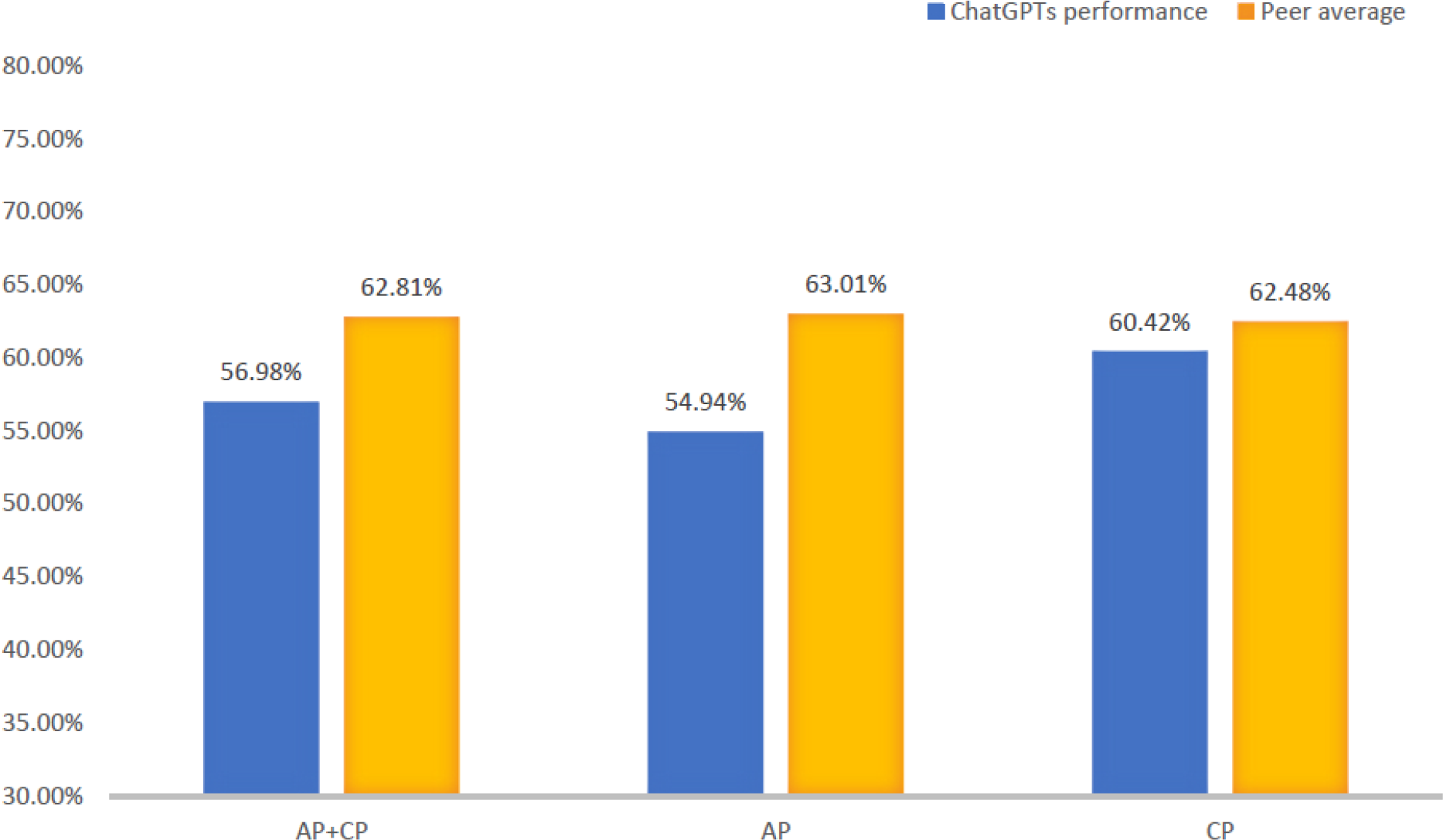
ChatGPT’s Performance with Peer Average in ASCP Resident Question Bank 2022. AP: Anatomic pathology; CP: Clinical pathology

**Figure 4:**
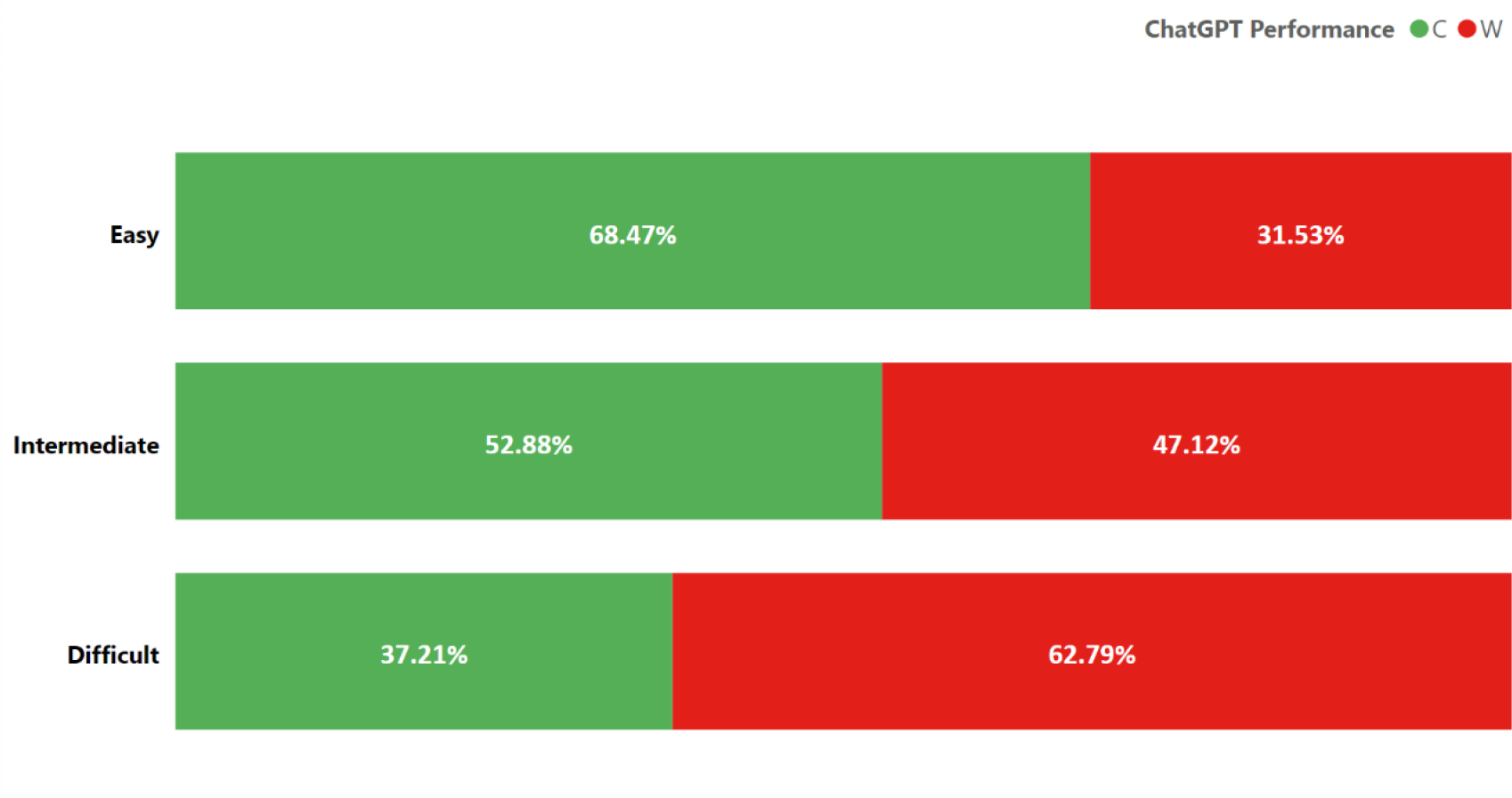
ChatGPTs performance based on difficulty level of questions C=Correct, W=Wrong.

There were 15 and 6 subcategories within the AP and CP sections respectively. In the AP section, ChatGPT’s correctness ranged from 30% (breast) to 71.4% (dermatology). It outperformed the peers in dermatology (71.43 % vs 47.04%), autopsy (60 % vs 54.64%), and gynecologic pathology (66.67% vs 63.33%). Its performance tallied with the peers in forensics (55.56%) and molecular pathology (70%) (Table 2 and Figure 5).

**Figure 5:**
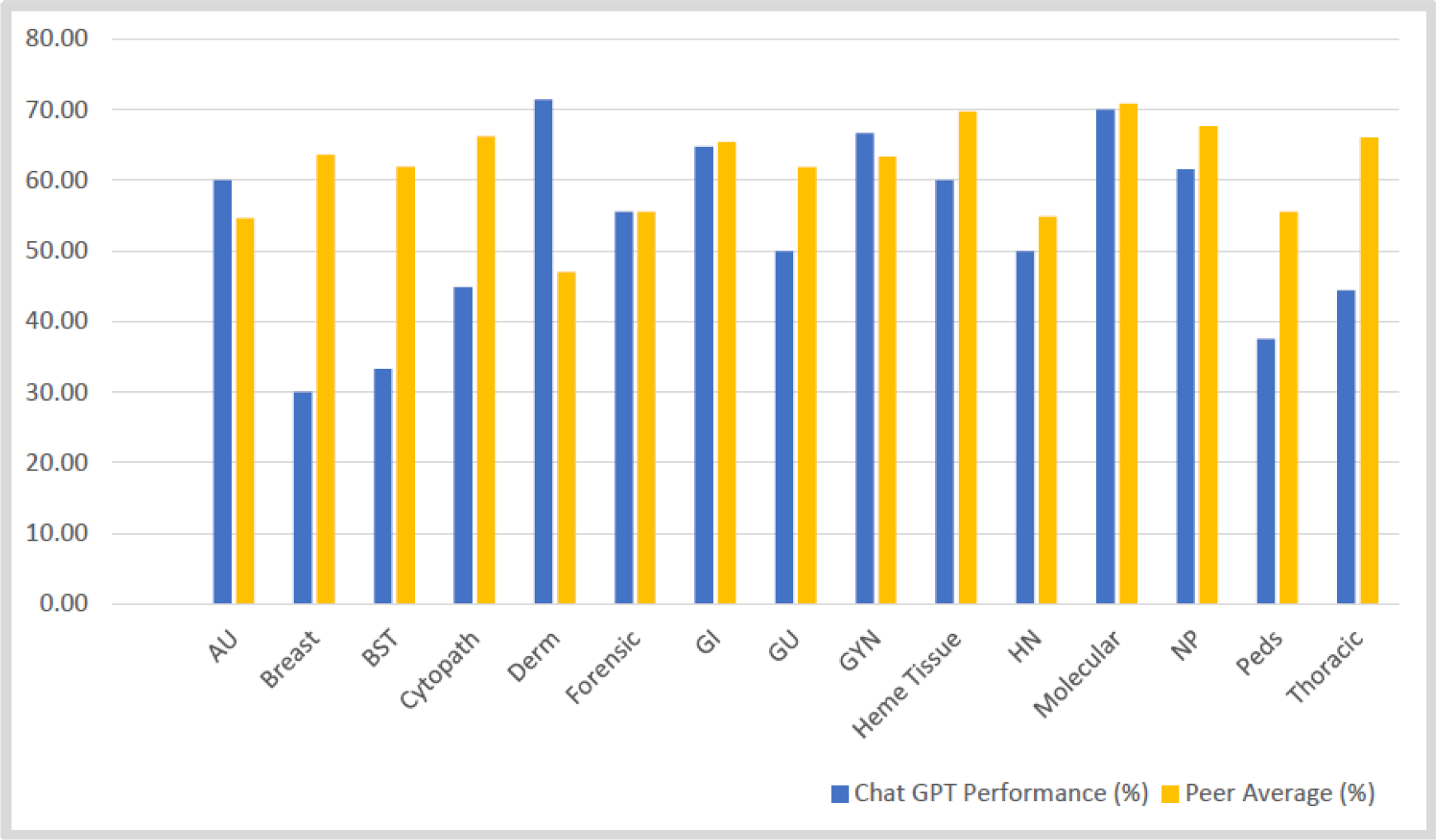
Comparison of ChatGPT’s performance in Anatomic Pathology with Peer Average. AU: Autopsy; BST: Bone and soft tissue; Cytopath: Cytopathology; Derm: Dermatology; GI: Gastroenterology; GU: Genitourinary; GYN: Gynecologic; Heme: Hematology; HN: Head and neck; NP:Neuropathology; Peds: Pediatric

In the CP section ChatGPT’s correctness was 100% in immunology, however, only one question was tested. ChatGPT’s performance after excluding immunology ranged from 71.43 % (hematology) to 50% (microbiology). It outperformed the peers in coagulation (70.83% vs 69.04%), hematology (71.43% vs 67.21%), and chemistry (56.52% vs 51.62%) (Table 3 and Figure 6).

**Figure 6:**
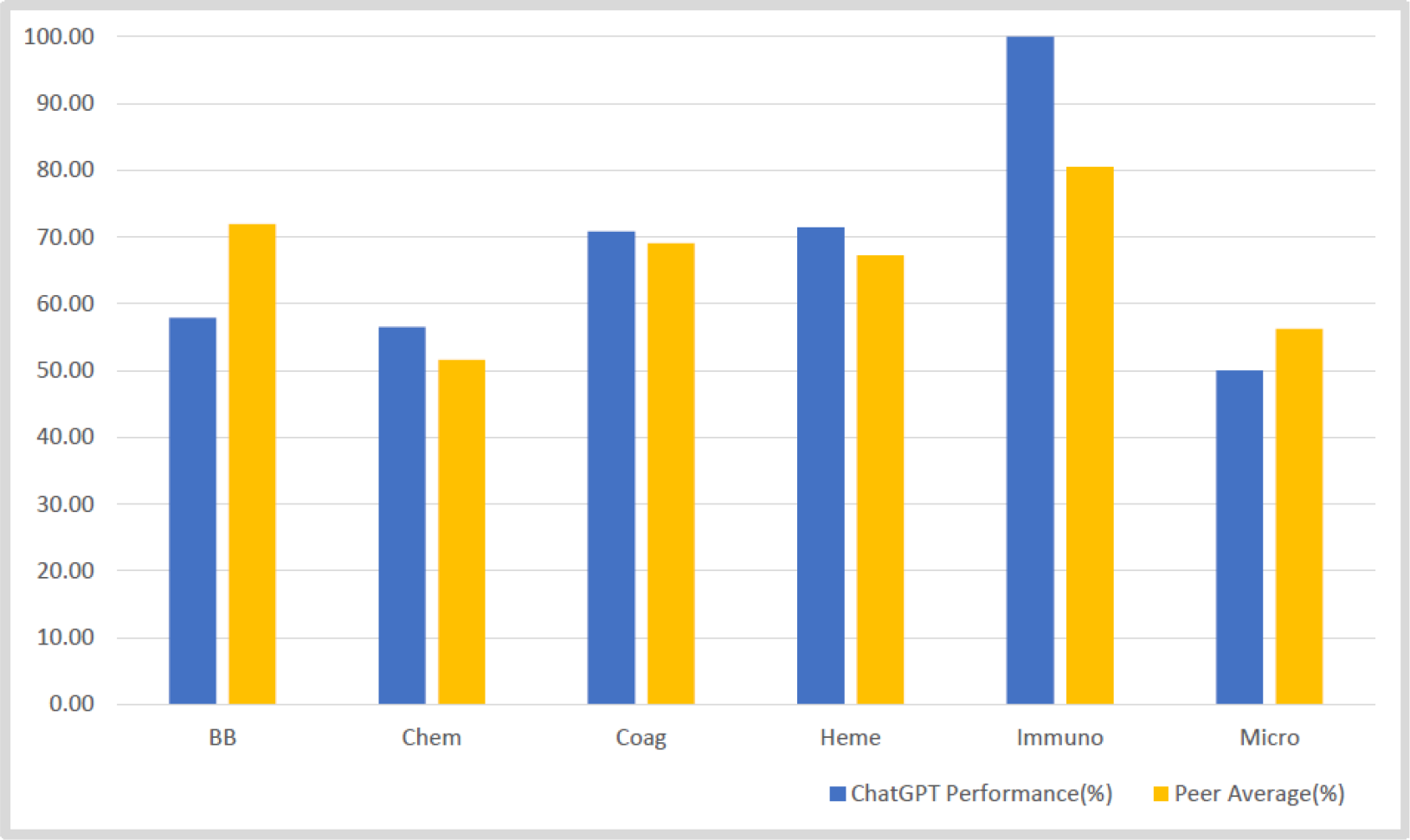
Comparison of ChatGPT’s performance in Clinical Pathology with Peer Average. BB: Blood bank; Chem: Chemistry; Coag: Coagulation; Heme: Hematology; Immuno: Immunology; Micro: Microbiology

The overall performance of Chat GPT on the second run was comparable to its first run (56.2% vs 56.97%) (Table 4). Upon analyzing each subspecialty individually, it was observed that in the majority of the fields its performance on the second run was better or on par with the first run. However, in a few disciplines such as forensics, hematology, molecular, neurology, chemistry, immunology, and microbiology, its performance exhibited a marginal decline, with the second run yielding one less correct response in most of these disciplines.

## Discussion

The extensive range of knowledge and unique interactive capabilities possessed by ChatGPT enables it to be utilized in various innovative ways, such as emulating a teacher, acting as a tool for generating ideas and receiving feedback, or even serving as a virtual study companion.^10^ However, for these applications to be truly effective, it is crucial for ChatGPT to exhibit performance that is equal to or surpasses that of a human in terms of knowledge and assessment.

We utilized ASCPs RQB-2022, a study tool used by pathology residents for continued education and preparation for their Pathology Board Examinations to evaluate ChatGPT’s pathology knowledge. Our study shows that the pathology knowledge of ChatGPT is not comprehensive, deviating from the anticipated level of proficiency characteristic of an AI tool. Moreover, its knowledge base falls considerably below that of pathology residents undergoing training. Additionally, our findings indicate that the tool’s pathology knowledge is rudimentary, as it demonstrated superior performance on easier questions compared to intermediate and difficult ones. Its performance exhibited a slight advantage in CP compared to AP. The overall difficulty level of CP questions tested were easier compared to the AP questions and this could possibly explain the reason for such results. Importantly, ChatGPT provided detailed explanations for all answer options and the rationale behind selecting the correct option for each question. While these explanations appeared logical and easily comprehensible, the accuracy of their content remains questionable based on the results obtained from this study.

Similar studies in testing ChatGPT’s medical knowledge have been conducted utilizing the United States Medical Licensing Examination (USMLE) question bank and American College of Gastroenterology (ACG) self-assessment tests. The findings from these studies have been inconsistent, as ChatGPT demonstrated proficiency and successfully passed the USMLE exam, demonstrating its potential in the field of medical education.^11^ On the other hand, it encountered significant challenges and could not meet the requirements to pass the ACG test.^12^ A potential explanation that could account for the varied outcomes observed may stem from disparities in the knowledge foundation upon which the examinations are constructed. USMLE encompasses a comprehensive evaluation of fundamental medical knowledge such as basic science and basic clinical vignettes. Conversely, ACG is a more specialized test, necessitating extensive expertise within a specific domain of medicine.

A unique aspect of this study was assessing the reproducibility of its performance by doing a second run. ChatGPT has the capacity for training through feedback. It can be fed with the correct response after providing an erroneous one, resulting in a correct answer during a subsequent iteration. To circumvent this training process, we deliberately abstained from providing ChatGPT with correct responses following its initial output. Though its overall performance on the second run was comparable to the first, its performance varied drastically in some of the subspecialty fields. This variability in its response could be multifactorial. ChatGPT is trained on a vast amount of text data, which introduces variability in the information it has learned. The model generates responses based on patterns and associations in the training data, and sometimes these patterns can result in different interpretations or perspectives on a given question.^13^Furthermore, it incorporates a degree of randomness during inference to enhance its diversity of responses. This randomization can introduce further variability, particularly when multiple plausible answers exist for a given question.^14^

Though ChatGPT has emerged to be a huge success, such results on medical education are due to the fact that it has not been trained on an extensive medical database. The field of medicine is dynamic, with new guidelines, and treatment options emerging regularly. ChatGPT’s responses are based on a static dataset, potentially limiting its knowledge and thereby the accuracy and relevance of its responses. Furthermore, the ability to comprehend the context of a question is crucial in the medical field, as it enables appropriate interpretation and decision-making. ChatGPT, although proficient in language processing, might have encountered challenges in grasping the subtle changes and clinical implications presented in the practice questions. Another possible factor is the inability of ChatGPT to draw upon real-world experiences, making it challenging to provide answers to clinical scenario-based questions.

In our study, we observed that ChatGPT exhibited superior performance compared to resident peers in specialized domains like dermatopathology, autopsy, gynecologic pathology, coagulation, hematology, and chemistry. This suggests that with additional training on an extensive medical dataset, it has the potential to significantly contribute to pathology education. However, one major constraint regarding its utilization in the context of pathology education lies in its lack of proficiency in image recognition, thereby impeding its ability to facilitate comprehension of the intricate microscopic characteristics exhibited by diverse disease entities. The incorporation of this functionality should be contemplated as a prospective enhancement in subsequent updates. A major limitation of our study resides in its descriptive nature. Majority of the subcategory sections contained a limited number of questions, which precluded our ability to perform meaningful statistical analysis.

## Conclusion

Pathology is an essential component of healthcare systems, the cornerstone of diagnosis and paves way for tailored disease management. With advancements in this field knowledge is ever growing. Being a field that has welcomed artificial intelligence such as digital pathology and incorporating it to day-to-day practice, the advent of language models, such as ChatGPT has sparked an interest in exploring its potential in supporting and enhancing pathology education. Our study found that though ChatGPT provides valuable pathology information, it cannot completely replace traditional educational resources or professional expertise. ChatGPT has the potential to be a more valuable resource if trained on an advanced and specialized medical dataset.

## Data Availability

All data produced in the present work are contained in the manuscript.

